# Homologous and heterologous antibodies to coronavirus 229E, NL63, OC43, HKU1, SARS, MERS and SARS-CoV-2 antigens in an age stratified cross-sectional serosurvey in a large tertiary hospital in The Netherlands

**DOI:** 10.1101/2020.08.21.20177857

**Authors:** Brenda M. Westerhuis, Erwin de Bruin, Felicity D. Chandler, Chris R. B. Ramakers, Nisreen M.A. Okba, Wentao Li, Herman Goossens, Menno D. de Jong, Berend Jan Bosch, Bart L. Haagmans, Pieter L. A. Fraaij, Reina S. Sikkema, Marion P.G. Koopmans

## Abstract

Understanding the coronavirus (CoV) antibody landscape in relation to disease and susceptibility is critical for modelling of steps in the next phase during the current covid-19 pandemic. In March 2020, during the first month of the epidemic in The Netherlands, we performed cross sectional studies at two time points amongst patients of the Erasmus Medical Centre in Rotterdam, to assess the presence of antibodies against seasonal human coronaviruses (OC43, 229E, NL63, HKU1), emerging zoonotic coronaviruses (SARS, MERS) and SARS-CoV-2 in nine different age groups. We observed minimal SARS-CoV-2 reactivity early March (0.7% of sera), increasing to 3.0%, four weeks later, suggesting probably undetected cases during this early phase of the epidemic. Antibody responses were mostly coronavirus species specific at young age, but possible cross-reactivity between human seasonal CoVs was observed with increasing age.

---

Dear editor,

A total of 1608 left-over plasma samples from patients of nine age categories were tested in one dilution, of which 879 were collected in early March 2020 (timepoint 1: T1), and 729 in early April 2020 (timepoint 2: T2; table 1). All plasma samples were tested against 19 different antigens from seven human coronaviruses (hCoVs) (OC43, 229E, NL63, HKU1, SARS, MERS and SARS-CoV-2). Recombinant spike proteins of the S1 subunit (all coronavirus subtypes), S ectodomain (all except 229E) and nucleocapsid proteins (OC43, 229E, NL63, HKU1, MERS and SARS) were included. The SARS-1 nucleocapsid protein has been used in the SARS-CoV-2 specific analyses as described previously(1).

**Table 1.** Seropositivity against SARS-CoV-2 antigens.

| **S1** | timepoint 1 |  |  | timepoint 2 |  |  |
| --- | --- | --- | --- | --- | --- | --- |
|  | total | positive | percentage | total | positive | percentage |
| 6-12 months | 41 | 0 | 0,0 | 17 | 0 | 0,0 |
| 1-2 years | 75 | 0 | 0,0 | 14 | 0 | 0,0 |
| 2-5 years | 42 | 0 | 0,0 | 24 | 0 | 0,0 |
| 5-10 years | 96 | 1 | 1,0 | 34 | 0 | 0,0 |
| 10-20 years | 131 | 0 | 0,0 | 139 | 1 | 0,7 |
| 20-40 years | 131 | 0 | 0,0 | 140 | 3 | 2,1 |
| 40-60 years | 134 | 1 | 0,7 | 140 | 5 | 3,6 |
| 60-80 years | 124 | 0 | 0,0 | 139 | 7 | 5,0 |
| 80+ years | 105 | 0 | 0,0 | 82 | 2 | 2,4 |
| total | 879 | 2 | 0,2 | 729 | 18 | 2,5 |

| **S ecto** | timepoint 1 |  |  | timepoint 2 |  |  |
| --- | --- | --- | --- | --- | --- | --- |
|  | total | positive | percentage | total | positive | percentage |
| 6-12 months | 41 | 0 | 0,0 | 17 | 0 | 0,0 |
| 1-2 years | 75 | 0 | 0,0 | 14 | 0 | 0,0 |
| 2-5 years | 42 | 0 | 0,0 | 24 | 0 | 0,0 |
| 5-10 years | 96 | 0 | 0,0 | 34 | 0 | 0,0 |
| 10-20 years | 131 | 0 | 0,0 | 139 | 1 | 0,7 |
| 20-40 years | 131 | 0 | 0,0 | 140 | 3 | 2,1 |
| 40-60 years | 134 | 2 | 1,5 | 140 | 5 | 3,6 |
| 60-80 years | 124 | 0 | 0,0 | 139 | 7 | 5,0 |
| 80+ years | 105 | 0 | 0,0 | 82 | 2 | 2,4 |
| total | 879 | 2 | 0,2 | 729 | 18 | 2,5 |

| **NP** | timepoint 1 |  |  | timepoint 2 |  |  |
| --- | --- | --- | --- | --- | --- | --- |
|  | total | positive | percentage | total | positive | percentage |
| 6-12 months | 41 | 0 | 0,0 | 17 | 0 | 0,0 |
| 1-2 years | 75 | 0 | 0,0 | 14 | 0 | 0,0 |
| 2-5 years | 42 | 0 | 0,0 | 24 | 0 | 0,0 |
| 5-10 years | 96 | 0 | 0,0 | 34 | 0 | 0,0 |
| 10-20 years | 131 | 0 | 0,0 | 139 | 1 | 0,7 |
| 20-40 years | 131 | 1 | 0,8 | 140 | 4 | 2,9 |
| 40-60 years | 134 | 1 | 0,7 | 140 | 5 | 3,6 |
| 60-80 years | 124 | 2 | 1,6 | 139 | 9 | 6,5 |
| 80+ years | 105 | 0 | 0,0 | 82 | 2 | 2,4 |
| total | 879 | 4 | 0,5 | 729 | 21 | 2,9 |

Table 1. Seropositivity against SARS-CoV-2 antigens
| **S1, Secto, NP** | timepoint 1 |  |  | timepoint 2 |  |  |
| --- | --- | --- | --- | --- | --- | --- |
|  | total | positive | percentage | total | positive | percentage |
| 6-12 months | 41 |  | 0,0 | 17 |  | 0,0 |
| 1-2 years | 75 |  | 0,0 | 14 |  | 0,0 |
| 2-5 years | 42 |  | 0,0 | 24 |  | 0,0 |
| 5-10 years | 96 |  | 0,0 | 34 |  | 0,0 |
| 10-20 years | 131 |  | 0,0 | 139 | 1 | 0,7 |
| 20-40 years | 131 |  | 0,0 | 140 | 2 | 1,4 |
| 40-60 years | 134 | 1 | 0,7 | 140 | 5 | 3,6 |
| 60-80 years | 124 |  | 0,0 | 139 | 7 | 5,0 |
| 80+ years | 105 |  | 0,0 | 82 | 2 | 2,4 |
| total | 879 | 1 | 0,1 | 729 | 17 | 2,3 |

Positivity rates for each of the SARS-CoV-2 antigens were calculated according to the cut-offs determined, based on 100% specificity (Table 1, Figure 1, Supplemental figure). SARS-CoV-2 specific antibodies (one dilution) were analyzed, for T1 and T2 separately. Despite the short interval between both sampling moments, a clear rise in SARS-CoV-2 seropositivity was observed. The prevalence of antibodies to any of the SARS-CoV-2 antigens increased from 0.7% (6/ 879) at T1 to 3.0% (22/729) at T2. When only taking into account plasma samples testing positive for all three antigens, the prevalence was 0.1% (1/879) at T1 and 2.3% (17/729) at T2. Most antibody positive sera were detected in age groups 40–60 and 60–80. None of the children below five years of age were seropositive (Tabel 1).

**Figure 1;.**
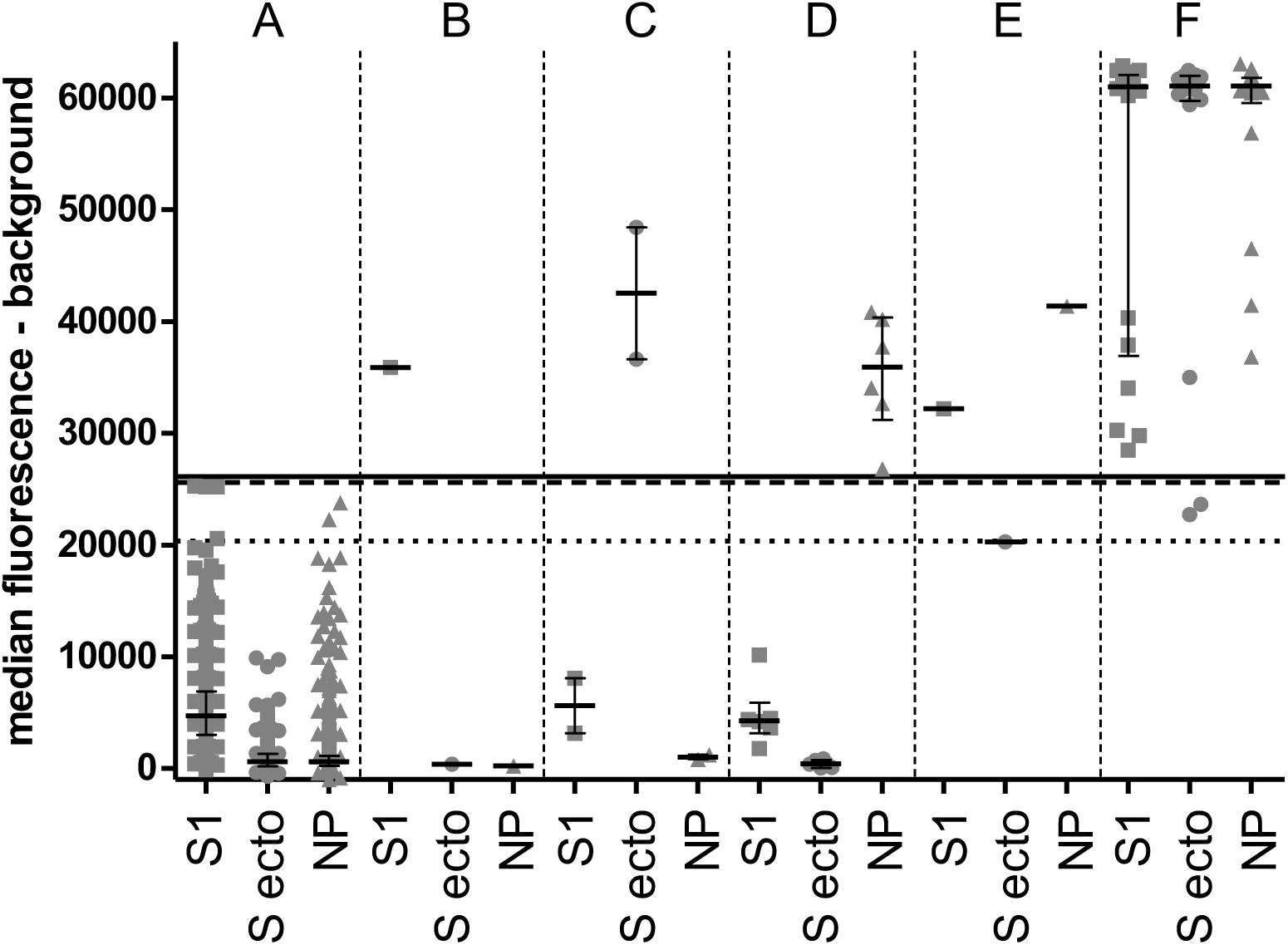
Overview of all fluorescent signals detected, depicting fluorescence on the Y-axis, and SARS-CoV-2 antigens on the X-axis grouped on positivity profile, where; negative for all antigens (A), only positive for S1 (B,n = 1), only positive for S-ecto (C, n = 2), only positive for NP (D, n = 6), positive for S1 and NP (E, n = 1) and positive for all 3 SARS-CoV-2 antigens (F, n = 18). Antigen specific cut-offs are depicted for S1 (horizontal line), S ecto (broken line) and NP (dotted line). Median values and interquartile range are depicted per group

In sera with SARS-CoV-2 responses, crossreactivity was shown against the SARS and MERS antigens (Figure 2). The majority of antibodies binding to antigens from seasonal human coronaviruses were present from an early age onwards in most plasma samples, although differences in antibody responses were detected between coronaviruses and between antigens from the same virus (Figure 2). The median fluorescence level of hCoV antibody reactivity was lowest in 6–24 month old children and increased from age groups 2–5 years old to > 20 years old, depending on the antigen. For some antigens a slight decrease in binding antibodies was observed in individuals above 80 years old.

**Figure 2;.**
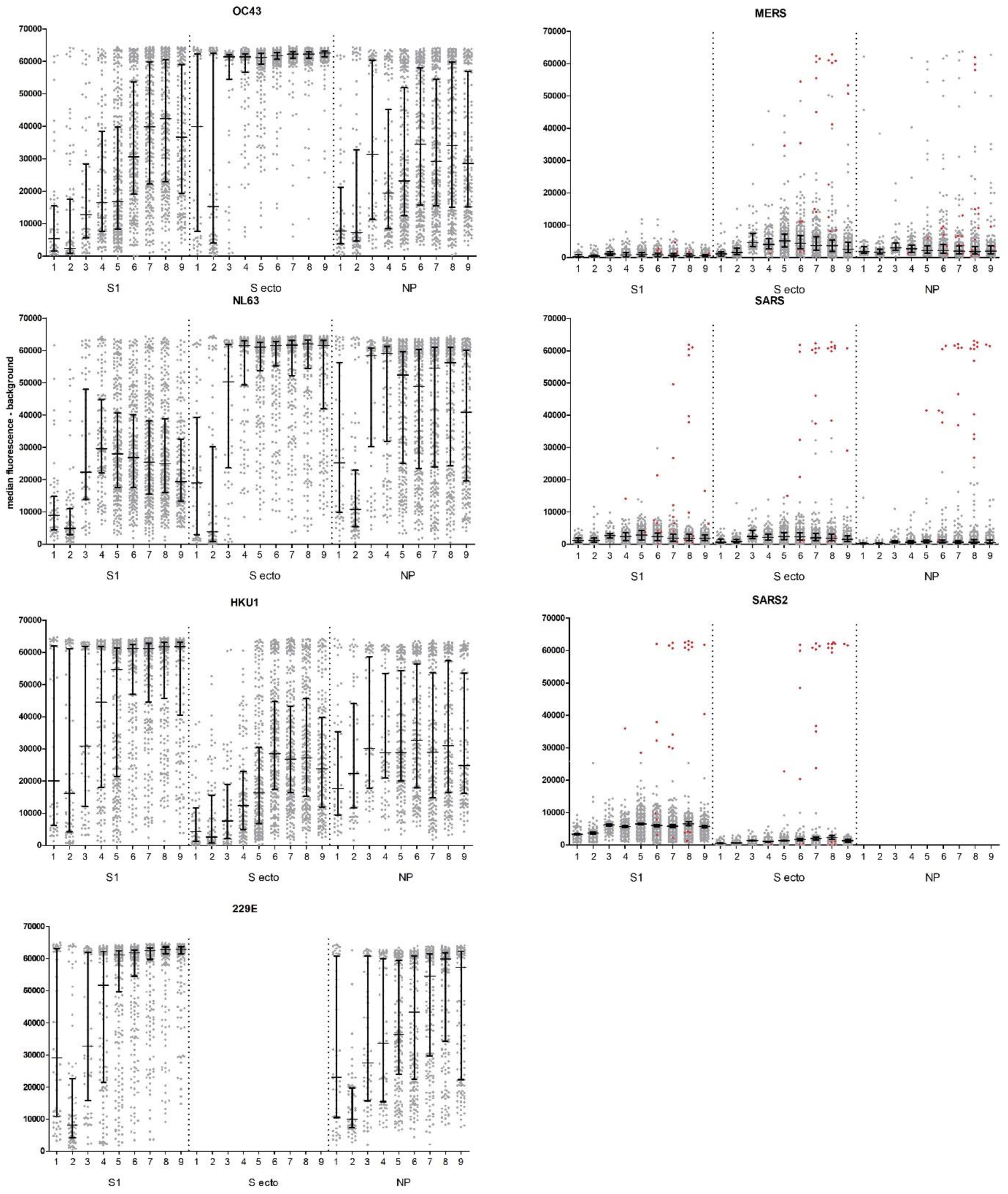
Fluorescence signals against CoV antigens. Sera with fluorescence signals above the cutoff against SARS-CoV-2 S1 and S-ecto and SARS-CoV NP are indicated in red.

To determine how reactivity of the different seasonal hCoV antibody titers correlate with (multiple) infections during lifetime, endpoint titers were calculated for all hCoV antigens for age groups 0–2y (n = 36 samples), 2–10y (n = 38), 10–40y (n = 37), 40+ (n = 58) and 60+ (n = 39). When looking at all data combined, highest correlations were observed between S1 and the S ectodomain from the same virus, and within alpha and beta coronaviruses, respectively. When breaking the groups down in age groups, mainly strain specific signals were observed in the lowest age group, with broadening of the correlation of antibody titers within the alpha and the betacoronaviruses between the ages of 2 and 40 years, and further broadening of responses between alpha and betacoronaviruses in individuals of 60 years and older (Figure 3).

**Figure 3;.**
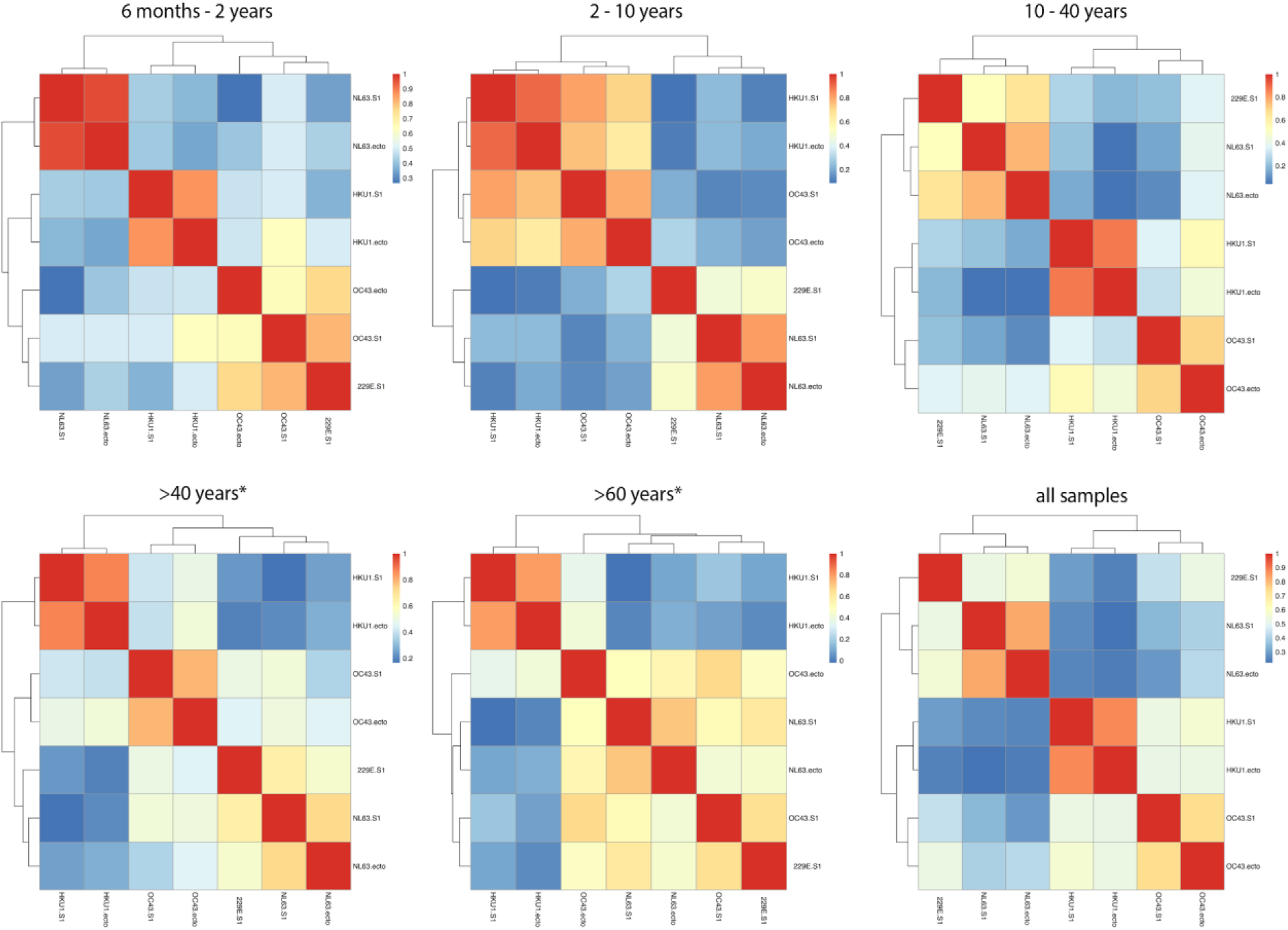
Pearson correlation heatmaps of binding antibody titers to HKU1, OC43, NL63 and 229E S1 and ectodomains

Serological assays have been shown to be of great value in population surveys of viral infections additional to virus detection assays. In this study we used a multiplex serological protein microarray to detect possible infections of SARS-CoV-2 early in the epidemic among the general population, including children and adolescents, as well as age stratified serological responses against a broad range of other CoV strains. The seropositivity at 4 weeks after confirmation of the first cases of SARS-CoV-2 in our study population revealed early, probably undetected, cases during this period, confirming the importance of adding serosurveys to assess the extent of spread of COVID-19. The highest seroprevalence was detected in the group 40 years and older.

Our data are in agreement with results of screening among human blood donors between 1–15 April, showing an average seroprevalence of 2.7% country wide, up to 9.5% in the hardest-hit areas (2). Worldwide most early seroprevalence studies showed similar numbers ranging from 0.4 – 14% (3), although seroprevalence can be considerably higher among outbreak settings (4, 5). Results of serological testing in a low prevalence situation need to be interpreted with caution, as low-level cross-reactivity has been observed for most currently available assays (6). A good practice can be to use combined assay results, counting only samples that had antibody reactivity to two or three of the SARS-CoV-2 antigens as positives(7). By using this criterion, and the high stringency cut-off defined by ROC analysis including serum samples form persons with recent seasonal coronavirus infections, we trust that our estimates are robust.

The use of leftover specimens across age ranges does provide a broader picture of community circulation than can be obtained with blood donors. The lack of SARS-CoV-2 antibody positive children below five years of age may be explained by lower infection rates or susceptibility in younger age groups, a lower proportion of severe cases, or both (8–10). We therefore were interested to assess whether the lack of SARS-CoV-2 infections was potentially related to immunity from other coronavirus infections.

The four endemic hCoVs (229E, NL63, OC43, and HKU) account for about 10% of all upper respiratory tract infections in adults (11, 12), but most CoV cases among healthcare seeking individuals with respiratory symptoms are detected in children < 5 years old and elderly > 64. In a limited number of clinical case series in Europe, OC43 and NL63 were most commonly detected, while HKU1 and 229E appear more infrequently or even in a temporal pattern (11, 13, 14). In our study, the prevalence and the magnitude of antibody positive signals increased from two years of age, although at a different rate for the different viruses. The antibodies were mostly specific for the different human CoVs in the younger age group, with limited correlation between signals for antigens from different human seasonal coronaviruses belonging to the same genera. Therefore, we find no evidence for presence of cross-reactive antibodies as an explanation for the seemingly lower rate of SARS-CoV-2 infections in children.

This pattern changed with increasing age, with increasing correlation between antibody signals for viruses within the same hCoV in adults, and even across hCoVs in older individuals. Cross reactivity of antibodies between seasonal coronaviruses and SARS have been described in the past. During an OC43 outbreak in British Colombia in 2003, cross-reactive antibodies to the SARS-CoV nucleocapsid protein have been shown (15) and a recent study showed very limited cross-reactivity with the SARS-CoV-2 ectodomain and S2 antigens in a naïve population, but not with the other SARS-CoV antigens (RBD, S1 and NP) (16). Our assay cut-off was set for high specificity of the SARS-CoV-2 measurements and therefore low-level binding would not be detected. However, we did observe cross-reactive antibodies to MERS-CoV S ectodomain and nucleocapsid antigens, without detectable responses to SARS-CoV-2, especially with increasing age. The possible broadening of anti S1 and S ectodomain antibodies with increasing age raises questions about the functionality of cross-reactive antibodies, although cross reaction with SARS-CoV-2 was not detected.

The methods we used have inherent disadvantages. In agreement with the general data protection regulation samples were collected in an anonymous manner, and as a consequence patients may have been included more than once. In addition, while the use of leftover serum collections is a proven method for fast deployment of sero-surveys, the study was hampered by the fact that during the first wave of the COVID-19 pandemic non-essential hospital visits were downscaled and individuals with mild respiratory disease strongly discouraged to visit health care providers. This especially affected hospital visits in the pediatric population as also reflected in our study with low numbers of serum samples for the pediatric population obtained on time point 2. It may therefore be that individuals with mild symptoms avoided the hospital and thus our study may underestimate the true sero-incidence in catchment area.

Nonetheless, we present an overview of the coronavirus antibody landscape in persons across all age groups, and show specific responses to SARS-CoV-2 in the same multiplex assay format. The presence of possible cross-hCoV and cross-subgroup binding antibodies with increasing age warrants further study in relation to disease outcomes.

## Material and Methods

### Plasma sample collection

This protocol was based on the principle employed during the influenza A/H1N1 2009 pandemic by Public Health England as a pragmatic approach to gathering essential baseline data for population exposure assessment during a rapidly evolving epidemic (17). Leftover lithium-heparin plasma samples submitted for general 24/7 clinical chemistry analyses at the Erasmus medical center were collected. Samples from patients in which a SARS-CoV-2 PCR was performed were excluded regardless of the result. Collection of plasma samples was done over a 5 days period at 2 time points, March 2nd and April 3rd. To allow for additional or repeat testing of archived samples if required clinically, left-over samples were retrieved after 4 to 7 days of cold storage (4C). The majority of samples from the children’s cohort (< 18 years) were collected through finger or heel stick (capillary blood). The vast majority of blood from the adult age cohorts were collected via venepuncture (venous blood). Only information regarding the age was collected from the sera, covering nine age groups (Table 1). The study was approved by the institutions medical ethical board and informed consent was waived as data were collected anonymized (MEC-2020–0158).

For assay validation, a panel of 60 serum samples from patients with confirmed seasonal coronavirus infections (OC43, 229E, HKU1 or NL63), collected before the COVID-19 outbreak were obtained from the MERMAIDS-ARI study. A panel of 56 sera from confirmed SARS-CoV-2 patients, collected at more than 10 days post onset of symptoms, were used as a positive control group (by universitair ziekenhuis Antwerp) (11). Positive control serum was made from a pool of confirmed positive COVID-19 patients.

### Laboratory procedures

All sera were tested against 19 different antigens from 7 different coronaviruses known to have infected humans in the past (OC43, 229E, NL63, HKU1, SARS, MERS and SARS-CoV-2). Recombinant spike proteins of the S1 subunit (all coronavirus subtypes) or the S ectodomain (all except 229E) were expressed in HEK293 cells as described before (1, 18). Nucleocapsid proteins produced in Escherichia coli (for OC43, 229E, NL63 and HKU1; Medix Biochemica, Finland) or produced in baculovirus and insect cells (MERS and SARS; Sino biological, China) were commercially obtained.

Recombinant proteins were printed using a non-contact printer (sciFlexarrayer SX, Scienion, Germany) on glass microscope slides coated with 64 pads of nitrocellulose creating 64 protein arrays per slide which were used to measure binding antibodies present in serum, as described before (19). Briefly, serum was diluted 1 in 20 in Blotto blocking buffer (Thermo Fisher Scientific Inc., Rockford, USA) containing 0,1 percent Surfact Amps (Thermo Fisher Scientific Inc.) and used in one dilution. In addition, 18–20 randomly selected serum samples per age group were tested in six dilutions to obtain full antibody titers to allow correlation plots. For this, randomly selected serum samples were titrated in 4-fold serum dilution steps? ranging from 1 in 20 to 20480. Titer values were calculated by curve fitting using variable slope, minimal fluorescence of 1000 and maximum fluorescence of 65535, as done before using Graphpad Prism (9). After serum incubation and washing, slides were incubated with goat anti human IgG (Fab-specific) conjugated to Alexa Fluor 647 (Jackson Immunoresearch Laboratories Inc., West Grove, USA) 1 in 1000 diluted in Blotto blocking buffer containing 0,1 percent Surfact Amps. Single spot median fluorescent signals using per spot background correction were measured using a Powerscanner (Tecan Group Ltd, Mannedorf, Switzerland).

## Analysis

Fluorescent cut-offs to determine positivity at a 1 in 20 serum dilution were determined, using the validation panels of seasonal CoV (negative group) and SARS-CoV-2 patients (positive group), by ROC-curves at which 100 percent specificity was chosen for high predictive value of positive signals, using Graphpad Prism (GraphPad software, version 5.01, San Diego, USA). Figures were made using Graphpad Prism (version 5.01) Correlation plots were made based on the fully titrated samples. Pearson correlation heatmaps were made using the pheatmap package using R Studio.

## Author Contributions

BW, EdB, RS and MK wrote the manuscript and executed the analyses, FC executed the laboratory analyses, CR, PF, MdJ, and HG were involved in the sampling design and sample collection, EdB, NO, BJB, WL, BH and MK were involved in assay design and validation, all authors read and contributed to the final manuscript.

## Data Availability

The datasets generated during and/or analysed during the current study are available from the corresponding author on reasonable request.

## References

1. Okba NMA, Müller MA, Li W, Wang C, GeurtsvanKessel CH, Corman VM, et al. Severe Acute Respiratory Syndrome Coronavirus 2-Specific Antibody Responses in Coronavirus Disease 2019 Patients. Emerg Infect Dis. 2020;26(7).

2. Slot E, Hogema BM, Reusken CBEM, Reimerink JH, Molier M, Karregat JHM, et al. Herd immunity is not a realistic exit strategy during a COVID-19 outbreak. 2020.

3. Bobrovitz N, Arora RK, Yan T, Rahim H, Duarte N, Boucher E, et al. Lessons from a rapid systematic review of early SARS-CoV-2 serosurveys. medRxiv. 2020:2020.05.10.20097451.

4. Fontanet A, Tondeur L, Madec Y, Grant R, Besombes C, Jolly N, et al. Cluster of COVID-19 in northern France: A retrospective closed cohort study. medRxiv. 2020:2020.04.18.20071134.

5. Houlihan C, Vora N, Byrne T, Lewer D, Heaney J, Moore DA, et al. SARS-CoV-2 virus and antibodies in front-line Health Care Workers in an acute hospital in London: preliminary results from a longitudinal study. medRxiv. 2020:2020.06.08.20120584.

6. GeurtsvanKessel CH, Okba NMA, Igloi Z, Bogers S, Embregts CWE, Laksono BM, et al. An evaluation of COVID-19 serological assays informs future diagnostics and exposure assessment. Nature Communications. 2020;11(1):3436.

7. den Hartog G, Schepp RM, Kuijer M, GeurtsvanKessel C, van Beek J, Rots N, et al. SARS-CoV-2-specific antibody detection for sero-epidemiology: a multiplex analysis approach accounting for accurate seroprevalence. medRxiv. 2020:2020.06.18.20133660.

8. Davies NG, Klepac P, Liu Y, Prem K, Jit M, Pearson CAB, et al. Age-dependent effects in the transmission and control of COVID-19 epidemics. Nature Medicine. 2020.

9. Lu X, Zhang L, Du H, Zhang J, Li YY, Qu J, et al. SARS-CoV-2 Infection in Children. N Engl J Med. 2020;382(17):1663–5.

10. Wu Z, McGoogan JM. Characteristics of and Important Lessons From the Coronavirus Disease 2019 (COVID-19) Outbreak in China: Summary of a Report of 72–314 Cases From the Chinese Center for Disease Control and Prevention. Jama. 2020.

11. Nickbakhsh S, Ho A, Marques DFP, McMenamin J, Gunson RN, Murcia PR. Epidemiology of Seasonal Coronaviruses: Establishing the Context for the Emergence of Coronavirus Disease 2019. J Infect Dis. 2020;222(1):17–25.

12. Paules CI, Marston HD, Fauci AS. Coronavirus Infections—More Than Just the Common Cold. Jama. 2020;323(8):707–8.

13. Heimdal I, Moe N, Krokstad S, Christensen A, Skanke LH, Nordbø SA, et al. Human Coronavirus in Hospitalized Children With Respiratory Tract Infections: A 9-Year Population-Based Study From Norway. J Infect Dis. 2019;219(8):1198–206.

14. Dijkman R, Jebbink MF, Gaunt E, Rossen JWA, Templeton KE, Kuijpers TW, et al. The dominance of human coronavirus OC43 and NL63 infections in infants. Journal of Clinical Virology. 2012;53(2):135–9.

15. Patrick DM, Petric M, Skowronski DM, Guasparini R, Booth TF, Krajden M, et al. An Outbreak of Human Coronavirus OC43 Infection and Serological Cross-reactivity with SARS Coronavirus. Can J Infect Dis Med Microbiol. 2006;17(6):330–6.

16. de Assis RR, Jain A, Nakajima R, Jasinskas A, Felgner J, Obiero JM, et al. Analysis of SARS-CoV-2 Antibodies in COVID-19 Convalescent Blood using a Coronavirus Antigen Microarray. bioRxiv. 2020:2020.04.15.043364.

17. Miller E, Hoschler K, Hardelid P, Stanford E, Andrews N, Zambon M. Incidence of 2009 pandemic influenza A H1N1 infection in England: a cross-sectional serological study. Lancet. 2010;375(9720):1100–8.

18. Wang C, Li W, Drabek D, Okba NMA, van Haperen R, Osterhaus A, et al. A human monoclonal antibody blocking SARS-CoV-2 infection. Nat Commun. 2020;11(1):2251.

19. Reusken CB, Haagmans BL, Muller MA, Gutierrez C, Godeke GJ, Meyer B, et al. Middle East respiratory syndrome coronavirus neutralising serum antibodies in dromedary camels: a comparative serological study. Lancet Infect Dis. 2013;13(10):859–66.

